# Clinical interpretation of machine learning models for prediction of diabetic complications using electronic health records

**DOI:** 10.1101/2022.03.11.22272039

**Authors:** Amanda Momenzadeh, Ali Shamsa, Jesse G. Meyer

## Abstract

Type 2 diabetes is a massive public health issue that continues to grow. The rate of diabetic complication progression varies across individuals. Understanding factors that alter the rate of complication onset may uncover new clinical interventions and help prioritize individuals for more aggressive management. Here, we explore how various machine learning models and types of electronic health records can predict fast versus slow diabetic complication onset using only patient data prior to diabetes diagnosis. We find that optimized random forests generally perform best among the tested models and combining all data sources yields the best predictive performance. A key differentiator of our study is our model interpretation, which identifies specific patient metrics from each dataset that play a unique role in the progression of each complication. Overall, our clinical interpretation of machine learning models can identify patients at risk for poorer outcomes years in advance of their diabetic complication.

## Introduction

According to the 2020 National Diabetes Statistics Report, an estimated 34 million (or 13%) of the United States (US) adult population has diabetes[1], and the prevalence of diagnosed diabetes among US adults is projected to rise to 61 million (or 18%) by the year 2060[2]. Diabetes is the most expensive chronic condition in the US; one of every four US health care dollars is spent on care for people with diabetes[3]. Globally, the direct health expenditure on diabetes in 2019 was $760 billion, which is projected to rise to $845 billion in 2045, with the largest expenditure in individuals 60-69 years old[4]. The prevalence of diabetes is highest among adults over 65 years, and the expected rise in diabetes is partially due to a decline in mortality in the diabetes population[2].

Long-term complications of diabetes are categorized as either microvascular, including nephropathy, neuropathy, and retinopathy, or macrovascular, including cardiovascular and peripheral vascular disease. Diabetes is the leading cause of new cases of blindness and kidney failure in the US, and was the 7^th^ leading cause of death in 2017[5]. There is also a growing list of newly recognized complications causally linked to diabetes, namely cancers, dementia, infections and liver disease[6]. The development of complications is influenced by various risk factors, including chronic hyperglycemia, obesity, dyslipidemia, hypertension, inflammatory cytokines, and altered miRNA expression causing accumulation of excess extracellular matrix in organs affected by diabetes[7]. Targeted therapies that delay or inhibit progression of diabetic complications are lacking and there remains a need for a better understanding of the pathophysiology underlying diabetic complications[8].

Maintaining blood glucose, blood pressure, and cholesterol levels within therapeutic goals is critical to reducing the risk of diabetic-related complications[9][10][11][12][13][14][15][16]. For example, every percentage point reduction in glycosylated hemoglobin(HgbA1c) can reduce the risk for microvascular complications by 40%[16]. However, twenty-one percent of US adults with diabetes who met laboratory criteria for diabetes were unaware of or did not report having diabetes[1], thus type 2 diabetes mellitus(T2DM) is often undiagnosed until irreversible diabetic complications have developed[9][16]. If detected and treated early, as much as 90% of blindness due to diabetic retinopathy may be preventable[16]. More accurate identification of individuals with T2DM at risk for complications would allow clinicians the chance for early intervention, leading to improved health outcomes for patients.

Electronic health records (EHR) are a powerful tool in understanding trends in disease development and creating prediction models that allow early interventions or modification of treatment options to improve patient outcomes. With the increasing use of EHR, large-scale patient data has become more accessible[17]. Machine learning (ML) has been a powerful tool aiding in clinical decision-making, identification of patients at risk for diseases (e.g., septic shock[18]), as well as repurposing of drugs for new indications[19]. As opposed to past algorithms built using a limited number of patient attributes (e.g., age, sex, smoking status, cardiovascular disease)[20], a ML model will learn the attributes with the most importance, allowing for identification of previously unknown risk factors of disease development. ML algorithms can be trained using a set of patient attributes (or features) and health outcomes given a clinical scenario, and then used to predict outcomes when provided previously unseen patient profiles.

A number of ML studies have investigated the development of diabetic-related complications using EHR data. Designing a model intended for use in the real-world clinical setting warrants evaluation of whether the model in fact learned what the user had intended. For example, how do the important features learned by the model align with established risk factors[21]? However, most ML studies focus on predictive performance and rarely provide meaningful explanation of their models[22], that is, patient characteristics that led to the prediction[23]. Model interpretation is arguably as or more important than model performance metrics[21]. Due to overwhelming evidence indicting poor reproducibility and reporting of clinical ML models, a 2020 paper made several recommendations for transparent and comprehensible reporting of results from ML studies that are directed at clinical researchers[24]. They recommend presenting high impact predictors of the model in a summary/tabular format and a narrative focusing on these variables[24]. Additionally, authors should discuss clinical interpretation of these variables with respect to the model outputs, including potential for translation to health care[24].

SHAP(SHapley Additive exPlanations) is a popular and effective approach published in 2017 for understanding each features’ contribution to a model’s predictions[21]. SHAP is unique in that it provides insights into the magnitude of importance for a feature as well as the direction a feature shifts a predicted outcome. Of six studies published after 2017 using ML to predict one or more diabetic complications, only one study displayed feature importance results using SHAP[17].

Additionally, this study only summarized the mean absolute SHAP value per feature, thereby forgoing the opportunity to understand whether each feature generally increased or decreased a prediction. The five other studies did not present any analysis of feature importance [25][26][27][28][29].

In this paper, we describe a study using patient EHR data prior to diagnosis of T2DM to predict a binary outcome of fast versus slow onset of T2DM complications using ML. In other words, at the time a patient is diagnosed with T2DM, can we predict whether that individual will be diagnosed with a diabetic complication faster or slower than 50% of the study population? Our main objectives were to (1) compare the utility of different EHR data types, (2) compare different model architectures, and (3) focus on interpretation of the models. Our results indicate the different performance of each input dataset: vitals, demographics, international classification of diseases (ICD) codes, social history, and laboratory. The combination of all five datasets gave the best prediction. Through SHAP, we also identify the models’ top predictors, a series of unique patient markers that differentially affect the onset of a complication, and attempt to validate these findings with the existing literature. Lastly, we investigated the level of medical care received by both fast and slow complication onset groups as a potential opportunity for improved diabetes care. This knowledge can be leveraged to target individuals as early as their T2DM diagnosis who are at risk for rapid onset of a diabetic complication.

## Results

### Patient characteristics

After filtering each data source for the last entry before or on the day of T2DM diagnosis, we selected 10,486 patients who had complete pre-diabetes data from all five EHR sources. Of these patients, 5,608 had nephropathy, 4,646 cardiovascular disease (CVD), 4,257 neuropathy, and 3,074 ocular disease (**Figure 1A**). A patient may have had multiple complications present within the study period.

**Figure 1.**
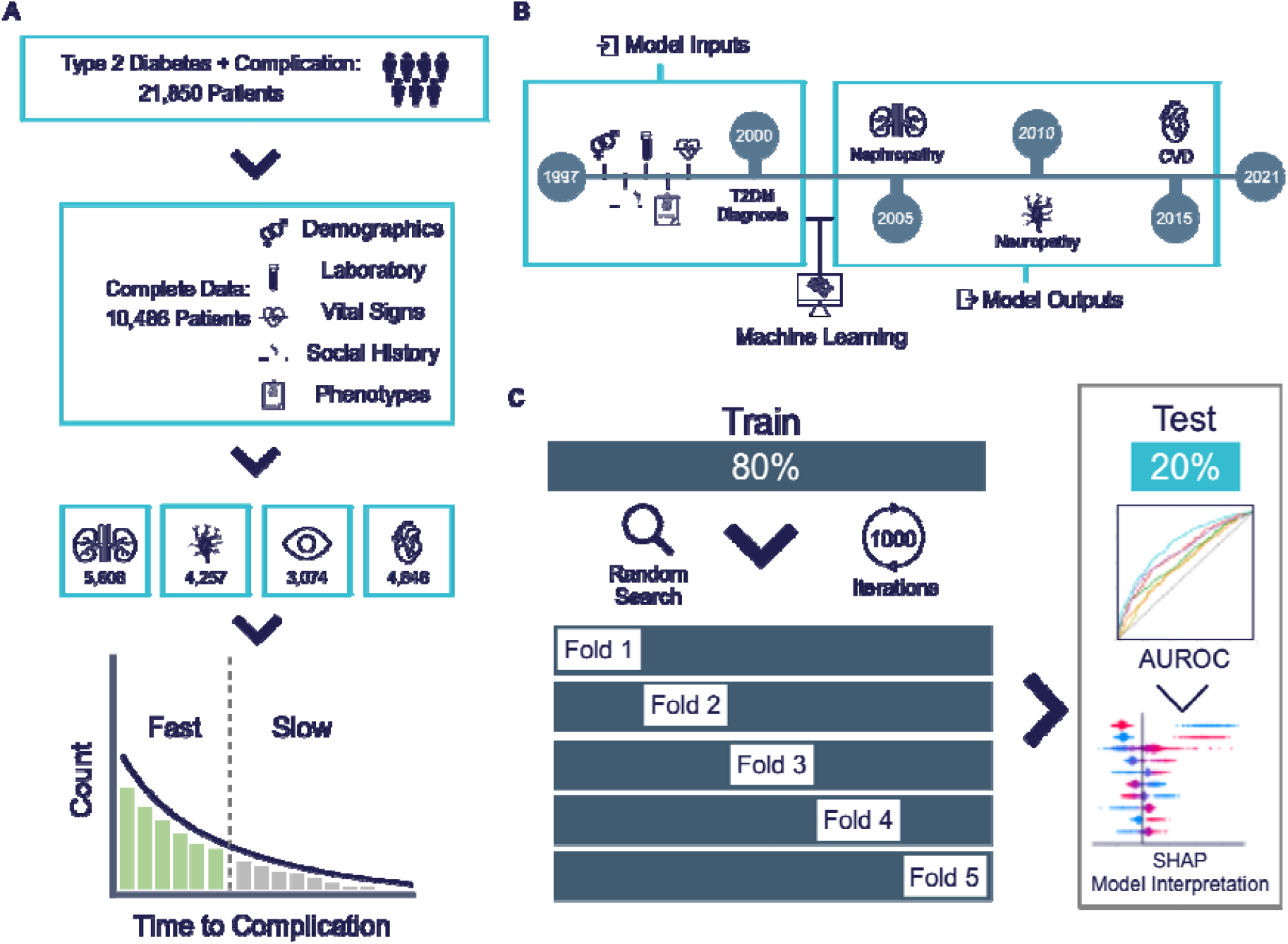
Flowchart depicting study development and analysis. **a**, Patients were selected from the Froedtert & MCW health network i2b2 de-identified patient database and multiple types of EHR were collected for each patient. Patients were divided into groups based on their diabetic complication, then further divided based on their rate of complication onset. **b**, Scheme showing the machine learning task concept with training inputs of EHR data and model outputs for an example patient. **c**, Scheme showing the machine learning model training strategy and the model evaluation with the test data.

Key patient characteristics were recorded in **Tables 1-4**. Across complications, patients in the slow complication onset group were diagnosed with T2DM at a younger age, were majority female, and had diabetes diagnosed for a longer duration compared to the fast complication onset group. The most prevalent race in both groups was Caucasian, followed by African American; the percentage of African Americans was consistently higher in the slow onset group. Across complications, BMI was higher in the slow onset group. Several patient risk factors for progression of diabetic complications were higher in the fast onset group across all complications, including percentage of patients with essential hypertension, hyperlipidemia, cigarette use, and known smokeless tobacco use status (although not always statistically significant). Glucose levels were significantly lower in the fast onset group across all complications, except neuropathy.

**Table 1.**
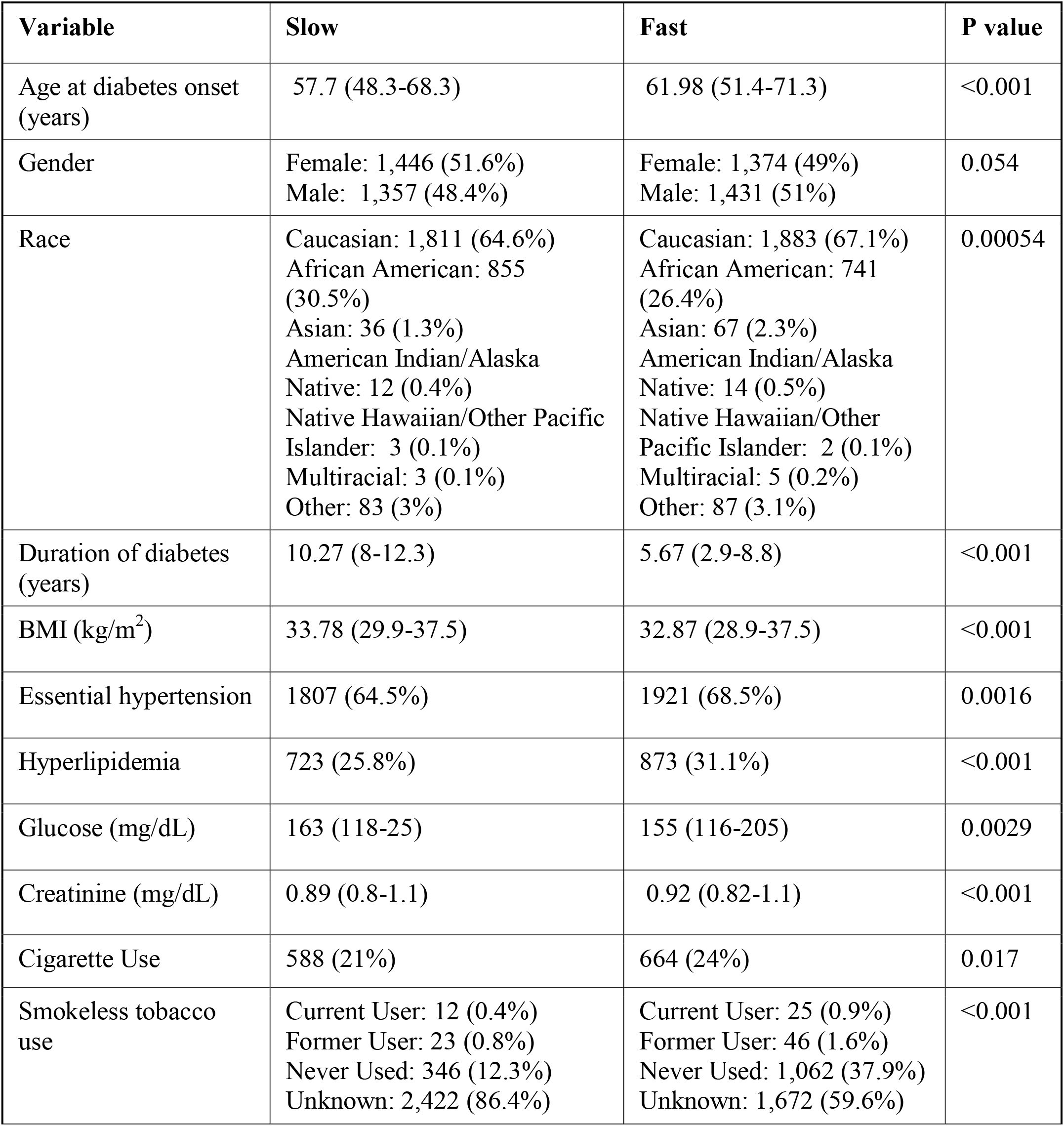
Baseline characteristics of patients who developed nephropathy during study period. Continuous variables are reported as median (inter-quartile range) and compared using a two-sided Mann-Whitney-U test. Categorical variables are reported as counts (percentage) and compared with the chi-square test. P-value≤0.05 was considered statistically significant.

| Variable | Slow | Fast | P value |
| --- | --- | --- | --- |
| Age at diabetes onset (years) | 57.7 (48.3-68.3) | 61.98 (51.4-71.3) | <0.001 |
| Gender | Female: 1,446 (51.6%)<br>Male: 1,357 (48.4%) | Female: 1,374 (49%)<br>Male: 1,431 (51%) | 0.054 |
| Race | Caucasian: 1,811 (64.6%)<br>African American: 855 (30.5%)<br>Asian: 36 (1.3%)<br>American Indian/Alaska Native: 12 (0.4%)<br>Native Hawaiian/Other Pacific Islander: 3 (0.1%)<br>Multiracial: 3 (0.1%)<br>Other: 83 (3%) | Caucasian: 1,883 (67.1%)<br>African American: 741 (26.4%)<br>Asian: 67 (2.3%)<br>American Indian/Alaska Native: 14 (0.5%)<br>Native Hawaiian/Other Pacific Islander: 2 (0.1%)<br>Multiracial: 5 (0.2%)<br>Other: 87 (3.1%) | 0.00054 |
| Duration of diabetes (years) | 10.27 (8-12.3) | 5.67 (2.9-8.8) | <0.001 |
| BMI (kg/m<sup>2</sup>) | 33.78 (29.9-37.5) | 32.87 (28.9-37.5) | <0.001 |
| Essential hypertension | 1807 (64.5%) | 1921 (68.5%) | 0.0016 |
| Hyperlipidemia | 723 (25.8%) | 873 (31.1%) | <0.001 |
| Glucose (mg/dL) | 163 (118-25) | 155 (116-205) | 0.0029 |
| Creatinine (mg/dL) | 0.89 (0.8-1.1) | 0.92 (0.82-1.1) | <0.001 |
| Cigarette Use | 588 (21%) | 664 (24%) | 0.017 |
| Smokeless tobacco use | Current User: 12 (0.4%)<br>Former User: 23 (0.8%)<br>Never Used: 346 (12.3%)<br>Unknown: 2,422 (86.4%) | Current User: 25 (0.9%)<br>Former User: 46 (1.6%)<br>Never Used: 1,062 (37.9%)<br>Unknown: 1,672 (59.6%) | <0.001 |

**Table 2.** Baseline characteristics of patients who developed neuropathy during study period. Continuous variables are reported as median (inter-quartile range) and compared using a two-sided Mann-Whitney-U test. Categorical variables are reported as counts (percentage) and compared with the chi-square test. P-value≤0.05 was considered statistically significant.

| Variable | Slow | Fast | P value |
| --- | --- | --- | --- |
| Age at diabetes onset (years) | 55.7 (45.6-65) | 58.9 (49-67.7) | <0.001 |
| Gender | Female: 1,138 (53.5%)<br>Male: 990 (46.5%) | Female: 1,090 (51.2%)<br>Male: 1,039 (48.8%) | 0.15 |
| Race | Caucasian: 1,345 (63.2%)<br>African American: 682 (32.1%)<br>Asian: 20 (0.9%)<br>American Indian/Alaska Native: 8 (0.4%)<br>Native Hawaiian/Other Pacific Islander: 0 (0%)<br>Multiracial: 3 (0.1%)<br>Other: 70 (3.3%) | Caucasian: 1,431 (67.2%)<br>African American: 568 (26.7%)<br>Asian: 38 (1.8%)<br>American Indian/Alaska Native: 7 (0.3%)<br>Native Hawaiian/Other Pacific Islander: 4 (0.2%)<br>Multiracial: 2 (0.1%)<br>Other: 79 (3.7%) | <0.001 |
| Duration of diabetes (years) | 10.7 (8.4-12.7) | 6 (3.5-8.6) | <0.001 |
| BMI (kg/m<sup>2</sup>) | 34.1 (30.3-38) | 33.6 (29.6-37.8) | 0.047 |
| Essential hypertension | 1274 (59.9%) | 1323 (62.1%) | 0.14 |
| Hyperlipidemia | 569 (26.7%) | 660 (31%) | 0.0024 |
| Glucose (mg/dL) | 160.5 (119-205) | 166.0 (121-209.7) | 0.038 |
| Creatinine (mg/dL) | 0.9 (0.8-1) | 0.9 (0.8-1) | 0.97 |
| Cigarette Use | 476 (22.4%) | 546 (25.7%) | 0.014 |
| Smokeless tobacco use | Current User: 10 (0.5%)<br>Former User: 14 (0.7%)<br>Never Used: 255 (12%)<br>Unknown: 1,849 (87%) | Current User: 19 (0.9%)<br>Former User: 40 (1.9%)<br>Never Used: 915 (43%)<br>Unknown: 1,155 (54.3%) | <0.001 |

**Table 3.**
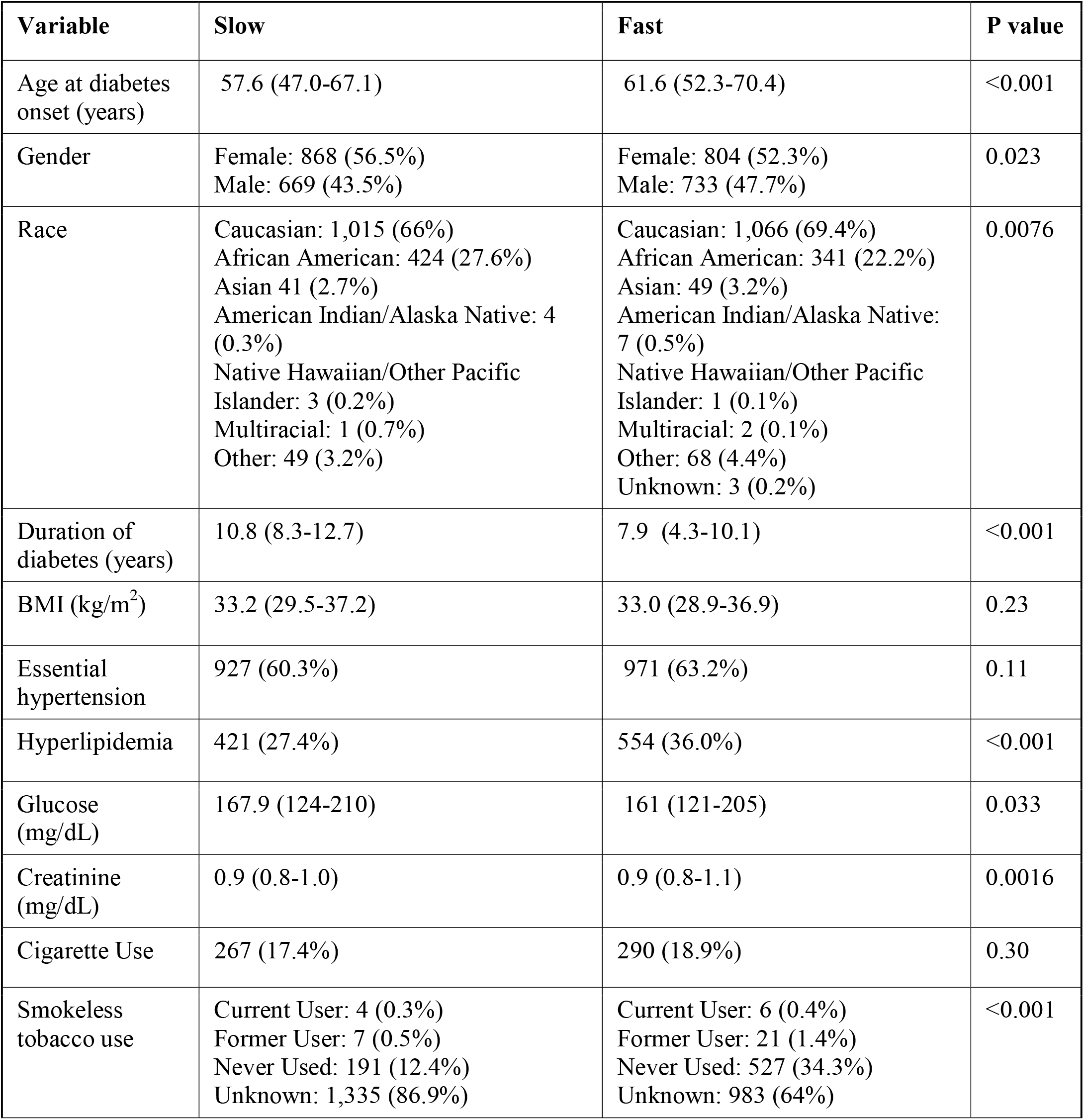
Baseline characteristics of patients who developed ocular disease during study period. Continuous variables are reported as median (inter-quartile range) and compared using a two-sided Mann-Whitney-U test. Categorical variables are reported as counts (percentage) and compared with the chi-square test. P-value≤0.05 was considered statistically significant.

**Table 4.** Baseline characteristics of patients who developed CVD during study period. Continuous variables are reported as median (inter-quartile range) and compared using a two-sided Mann-Whitney-U test. Categorical variables are reported as counts (percentage) and compared with the chi-square test. P-value≤0.05 was considered statistically significant.

| Variable | Slow | Fast | P value |
| --- | --- | --- | --- |
| Age at diabetes onset (years) | 59.43 (50.2-68.3) | 64.67 (55.3-73.4) | <0.001 |
| Gender | Female: 1,202 (51.8%)<br>Male: 1,119 (48.2%) | Female: 1,123 (48.3%)<br>Male: 1,202 (51.7%) | 0.019 |
| Race | Caucasian: 1,565 (67.4%)<br>African American: 656 (28.3%)<br>Asian: 26 (1.1%)<br>American Indian/Alaska Native: 7 (0.3%)<br>Native Hawaiian/Other Pacific Islander: 2 (0.1%)<br>Multiracial: 1 (0.04%)<br>Unknown: 1 (0.04%)<br>Other: 63 (2.7%) | Caucasian: 1,657 (71.3%)<br>African American: 537 (23.1%)<br>Asian: 40 (1.7%)<br>American Indian/Alaska Native: 13 (0.6%)<br>Native Hawaiian/Other Pacific Islander: 3 (0.1%)<br>Multiracial: 5 (0.2%)<br>Other: 69 (3.0%) | 0.0021 |
| Duration of diabetes (years) | 10.17 (7.8-12.4) | 5.91 (3.0-9.4) | <0.001 |
| BMI (kg/m<sup>2</sup>) | 33.8 (30.1-37.6) | 33.0 (29.1-37.2) | <0.001 |
| Essential hypertension | 1,536 (66.1%) | 1,589 (68.3%) | 0.12 |
| Hyperlipidemia | 646 (27.8%) | 697(30%) | 0.11 |
| Glucose (mg/dL) | 160 (118-205) | 157 (117-205) | 0.22 |
| Creatinine (mg/dL) | 0.86 (0.8-1) | 0.89 (0.8-1.1) | <0.001 |
| Cigarette Use | 525 (22.6%) | 570 (24.52%) | 0.14 |
| Smokeless tobacco use | Current User: 5 (0.2%)<br>Former User: 18 (0.8%)<br>Never Used: 376 (16.2%)<br>Unknown: 1,922 (82.8%) | Current User: 10 (0.4%)<br>Former User: 36 (1.6%)<br>Never Used: 778 (33.5%)<br>Unknown: 1,501 (64.6%) | <0.001 |

The histograms displayed in **Figure 2** show the distribution of times to complications and the two outcome groups. Patients within each complication group were divided into either fast or slow complication onset groups based on whether the time to complication was shorter or longer than the median time for that group, respectively. All four complications exhibit skewed distributions with a median of approximately 3 years. CVD had the shortest time to complication (2.95 years) and neuropathy had the longest time (3.26 years).

**Figure 2.**
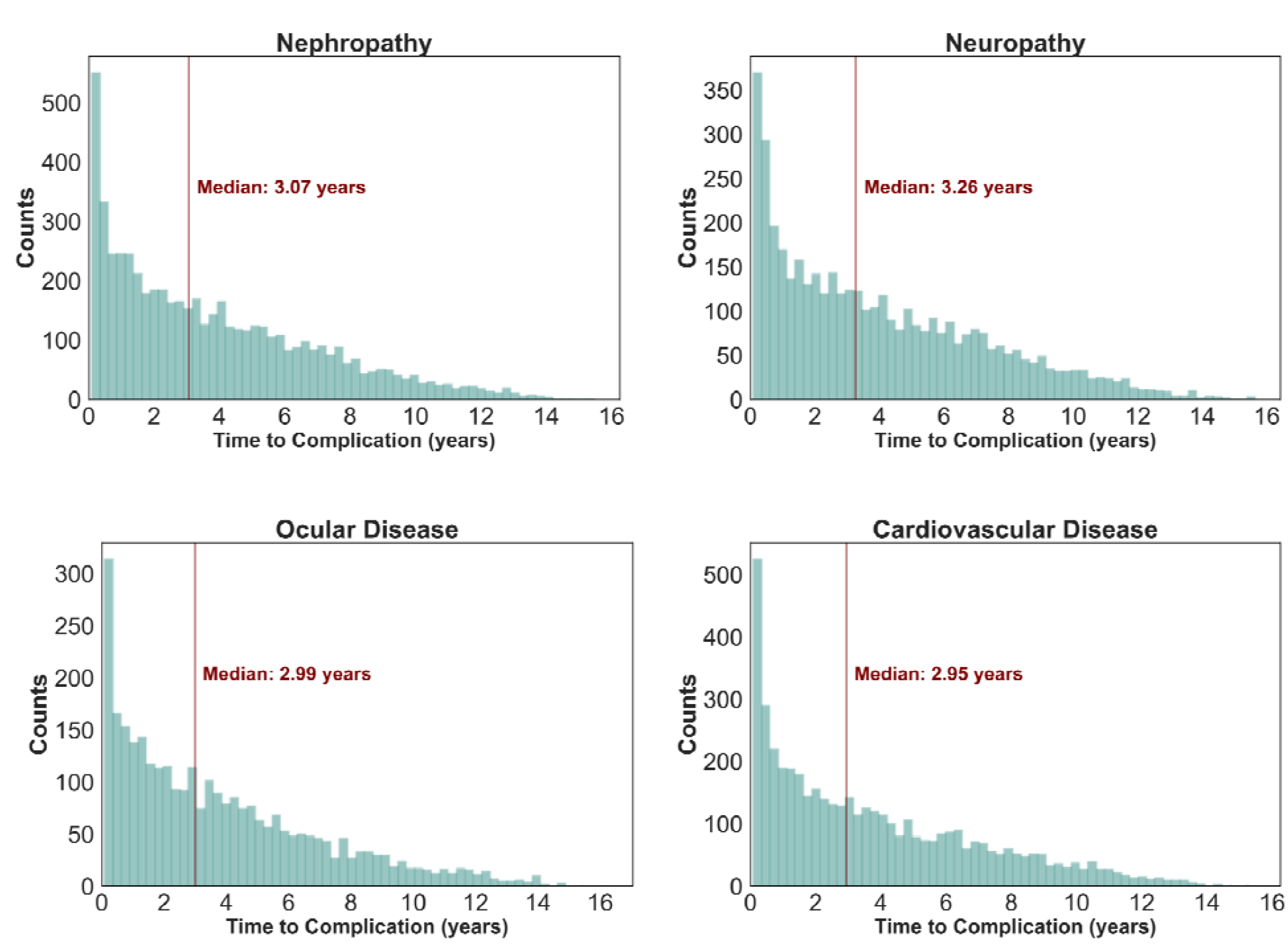
Time to complication histograms. Patient counts by time to diagnosis of complication (years) for individuals developing nephropathy, neuropathy, ocular disease, or CVD within the study. Red lines represent median time to complication diagnosis in each histogram.

### Model performance

To understand the relative utility of different types of EHR, the six different input datasets collected prior to the date of T2DM (phenotypes, demographics, vital signs, social-lifestyle history, laboratory data, and all inputs combined) were used to train one of six machine learning classification models (Gradient Boosting Decision Trees [GB], Support Vector Classification [SVC], Random Forest [RF], Extra Trees [ET], Logistic Regression [LR], and Adaptive Boosting [AdaBoost]) to predict one of four diabetic complications (**Figure 1B**). Data was split into 80% training and 20% test sets and models were optimized using a randomized search with 5-fold cross validation for up to 1,000 iterations using area under the receiver operating curve (AUROC) as the scoring metric. Models were refit with the training set data using the maximized hyperparameters obtained from the randomized search, and the test set was used to evaluate generalization performance of the best model (**Figure 1C**). AUROCs across each model and dataset combination is shown in **Table 5**. Using all datapoints combined as the model input, RF performed best in predicting nephropathy and neuropathy onset. ET and AdaBoost performed best in predicting CVD and ocular disease onset, respectively. Model calibration was assessed by plotting calibration curves of the observed versus predicted probabilities for the positive class across 10 evenly partitioned bins (**Supplementary Figure 1**). The brier scores for calibration plots were low, ranging from 0.204 to 0.223, indicating accurate probabilistic predictions.

**Table 5.** Test set AUROCs corresponding to each model input using six different ML models for each complication. Best AUROCs for each input (phenotypes, demographics, vitals, social-lifestyle history, laboratory, and all inputs combined) are bolded. SVC: Support Vector Classification, GB: Gradient Boosting Decision Trees, ET: Extra Trees, RF: Random Forest, AdaBoost: Adaptive Boosting, LR: Logistic Regression.

| Complication | Model | Phenotypes | Demographics | Vitals | Social | Labs | All |
| --- | --- | --- | --- | --- | --- | --- | --- |
| Nephropathy | SVC | 0.618 | 0.577 | 0.579 | 0.674 | 0.682 | 0.730 |
|  | GB | 0.615 | 0.581 | 0.585 | 0.677 | 0.661 | 0.736 |
|  | ET | **0.633** | 0.559 | 0.575 | **0.684** | 0.674 | 0.739 |
|  | RF | 0.625 | 0.577 | **0.593** | 0.670 | **0.684** | **0.747** |
|  | AdaBoost | 0.589 | **0.593** | 0.564 | 0.673 | 0.665 | 0.737 |
|  | LR | 0.612 | 0.580 | 0.567 | 0.671 | 0.672 | 0.728 |
| Neuropathy | SVC | 0.633 | 0.579 | 0.565 | 0.632 | 0.648 | 0.726 |
|  | GB | 0.629 | 0.582 | 0.559 | 0.666 | 0.666 | 0.713 |
|  | ET | 0.634 | 0.583 | 0.574 | **0.680** | 0.661 | 0.732 |
|  | RF | **0.638** | 0.582 | 0.578 | 0.674 | **0.671** | **0.737** |
|  | AdaBoost | 0.614 | **0.590** | **0.583** | 0.679 | 0.664 | 0.727 |
|  | LR | 0.624 | 0.583 | 0.524 | 0.677 | 0.645 | 0.724 |
| Ocular Disease | SVC | **0.567** | 0.608 | 0.507 | **0.633** | 0.662 | 0.649 |
|  | GB | 0.539 | 0.609 | 0.505 | 0.610 | 0.645 | 0.691 |
|  | ET | 0.530 | 0.601 | 0.520 | 0.632 | 0.656 | 0.682 |
|  | RF | 0.550 | 0.598 | **0.527** | 0.631 | 0.624 | 0.696 |
|  | AdaBoost | 0.502 | 0.612 | 0.508 | 0.599 | 0.586 | **0.707** |
|  | LR | 0.566 | **0.619** | 0.473 | 0.620 | **0.669** | 0.671 |
| CVD | SVC | 0.609 | 0.634 | 0.538 | 0.633 | **0.648** | 0.672 |
|  | GB | 0.603 | **0.637** | 0.516 | 0.606 | 0.634 | 0.694 |
|  | ET | 0.609 | 0.616 | **0.559** | **0.642** | 0.623 | **0.699** |
|  | RF | **0.613** | 0.623 | 0.542 | 0.599 | 0.626 | 0.693 |
|  | AdaBoost | 0.597 | 0.633 | 0.531 | 0.626 | 0.618 | 0.688 |
|  | LR | 0.603 | 0.632 | 0.539 | 0.619 | 0.647 | 0.679 |

Figure 3. displays overlayed AUROC plots for each complication with individual lines representing a different health dataset input. Across all complications, using all datasets combined as an input allowed for the highest model predictive performance compared to using individual datasets alone. Models were most effective in distinguishing between fast versus slow nephropathy onset (AUROC=0.75) and least effective in distinguishing between fast versus slow CVD onset (AUROC=0.70). Of the individual datasets, use of social history or laboratory values alone as inputs led to the highest model performance. Using vitals, demographics, or phenotypes alone led to poorer performance. Phenotypes outperformed vitals and demographics in prediction of nephropathy or neuropathy onset, however the demographics input was the strongest of these three inputs in predicting ocular disease or CVD onset.

**Figure 3.**
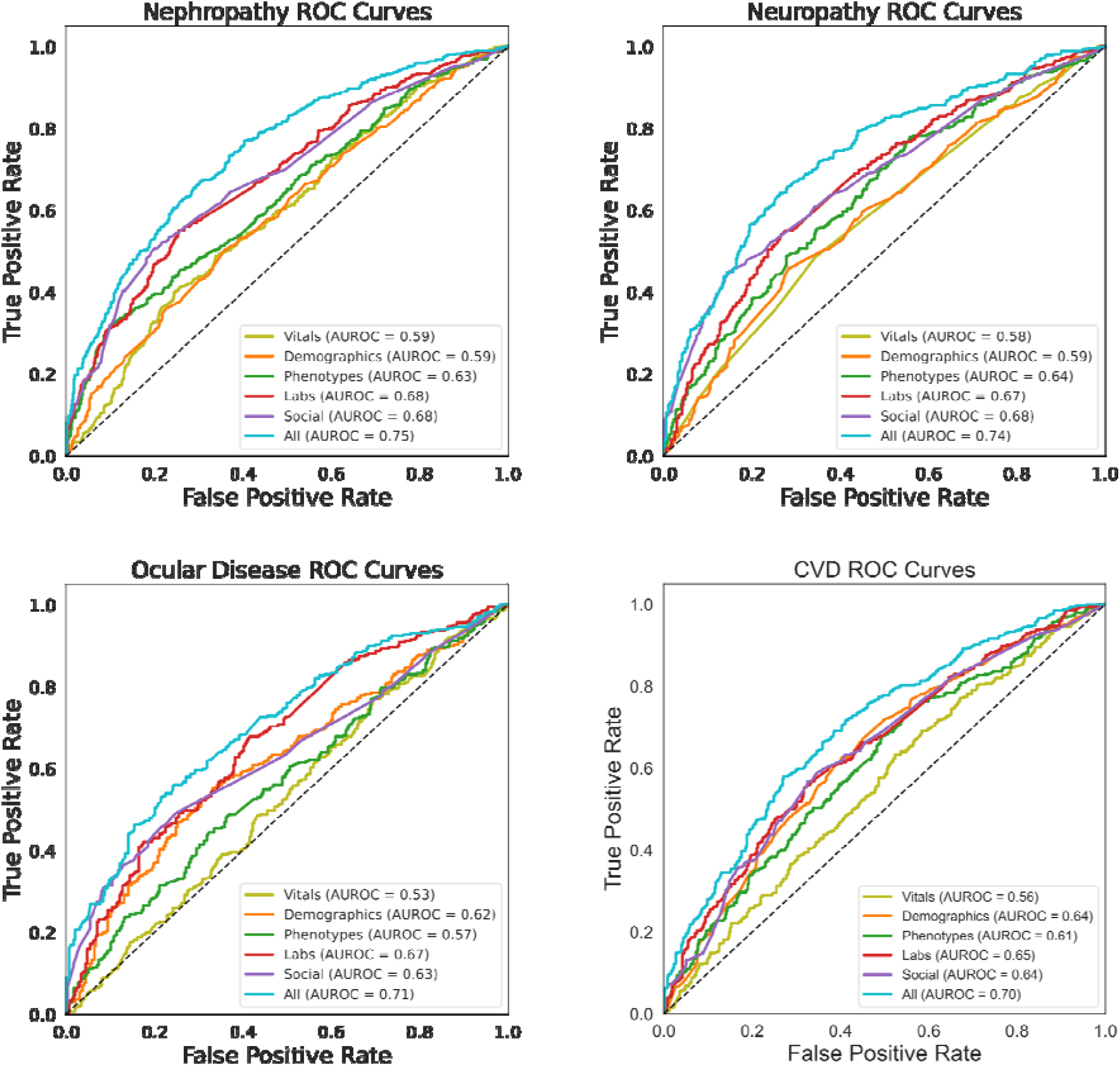
Overlayed area under receiver operating characteristic (AUROC) curves representing performance of each data source input for prediction of slow versus fast complication onset. AUROC’s corresponding to the best model are plotted for each input. AUROC of 0.5 (diagonal line) corresponds to a model that predicts the output with random chance and 1.0 corresponds to perfect classification. Data sources denoted in different colors: vitals (lime), demographics (orange), phenotypes (dark green), laboratory values (red), social history (purple), and all data sources combined (blue).

### Visualization of feature importance

SHAP was used to investigate how inputs to the model differentially affected the rate of diabetic complication onset (**Figure 4)**. As suggested by the AUROC curves, this analysis revealed that the models predominately leveraged social history and laboratory values in making predictions. The only demographic information that was a top 10 predictor was age at diabetes diagnosis. Phenotype was present only once within the top 10 predictors (i.e. hyperlipidemia in predicting ocular disease onset). Vitals, if present, tended to be of lower feature importance. Known smokeless tobacco use status, higher anion gap, and older age at diabetes diagnosis were associated with a faster onset across all four complications. A lower estimated glomerular filtration rate (eGFR) and higher mean platelet volume (MPV) were important in predicting fast onset of nephropathy, neuropathy and CVD, but did not play a role in prediction of ocular disease. Features unique to predicting fast ocular disease onset were a higher monocyte percentage, higher serum calcium, presence of hyperlipidemia and lower bilirubin. Lower mean corpuscular hemoglobin concentration (MCHC) and higher red cell distribution width (RDW) were associated with faster nephropathy onset. Extended SHAP plots displaying the top 25 predictors are presented in **Supplementary Figures 2-5**.

**Figure 4.**
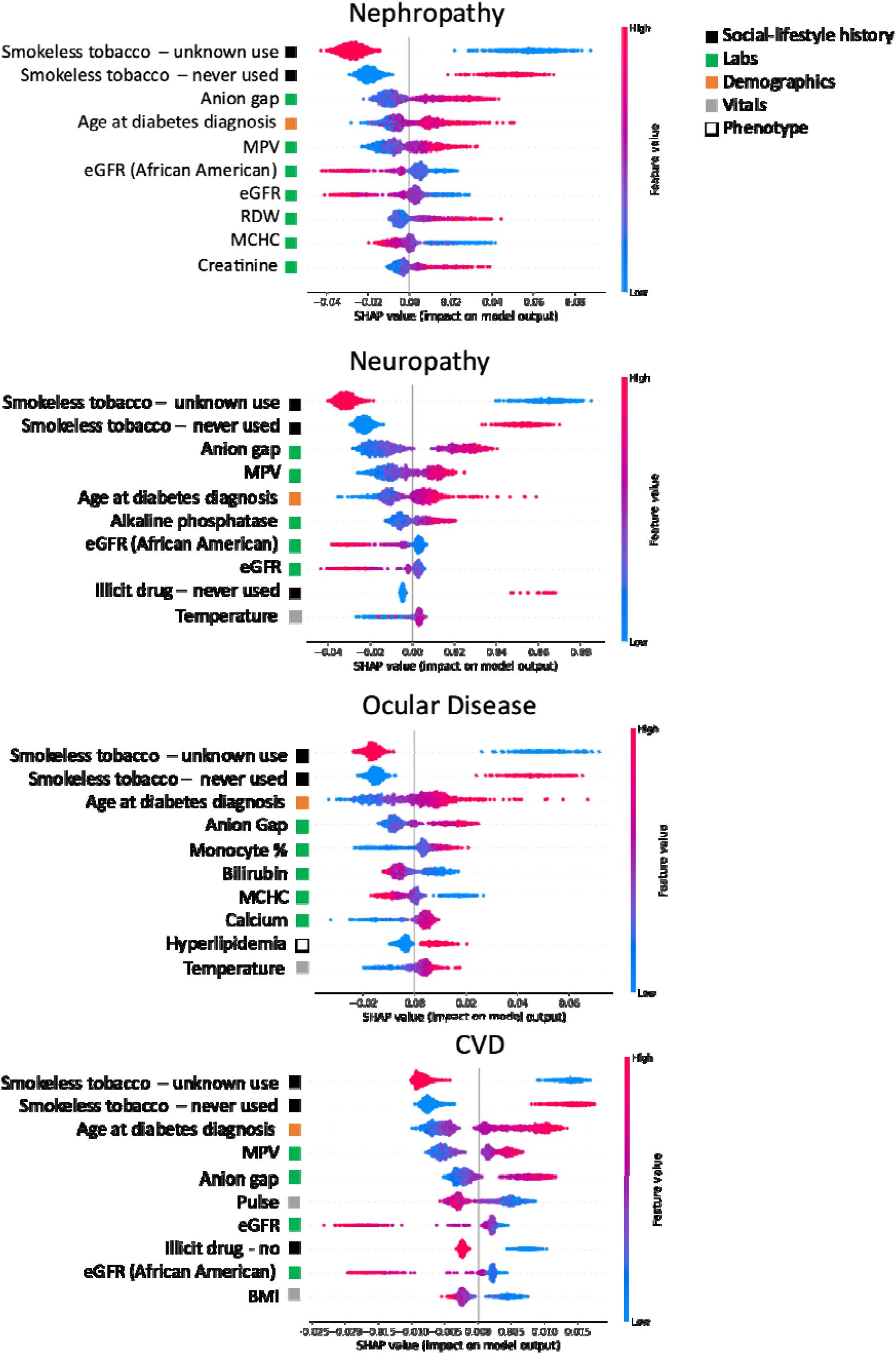
Top 10 features visualized using SHAP. Corresponding data source from which feature is derived is indicated by colored square. Individual patient contributions to the outcome are signified with red dots (high feature values), purple (intermediate), and blue (low). Y-axis represents importance of each feature. Dots with x values greater than and less than zero represent patients with a fast and slow complication onsets, respectively. MPV: mean platelet volume, eGFR: estimated glomerular filtration rate, RDW: red cell distribution width, MCHC: mean corpuscular hemoglobin concentration, BMI: body mass index.

### Medical care between T2DM and complication diagnosis

We further investigated patient engagement in medical care and types of visits sought between the time of T2DM and diabetic complication diagnoses between groups. We hoped to understand whether a lack of medical follow-up attributed to the faster development of complications. However, across all four complications, the fast complication onset group had significantly more medical visits per year **(Figure 5)**. Average median visits per year between time of T2DM and complication diagnoses across four complications was 27.3 in the fast onset group and 14.0 in the slow onset group. The most frequent types of visits recorded (**Figure 6**) were outpatient in nature (e.g. telephone, office visit, and therapy) compared to visits necessitating a higher level of care (e.g. emergency or inpatient hospital encounter). Taken in consideration with the observation that glucose levels were significantly lower in the fast onset nephropathy, ocular disease, and CVD groups, these findings potentially indicate that the fast onset group was engaged in routine, outpatient diabetes care.

**Figure 5.**
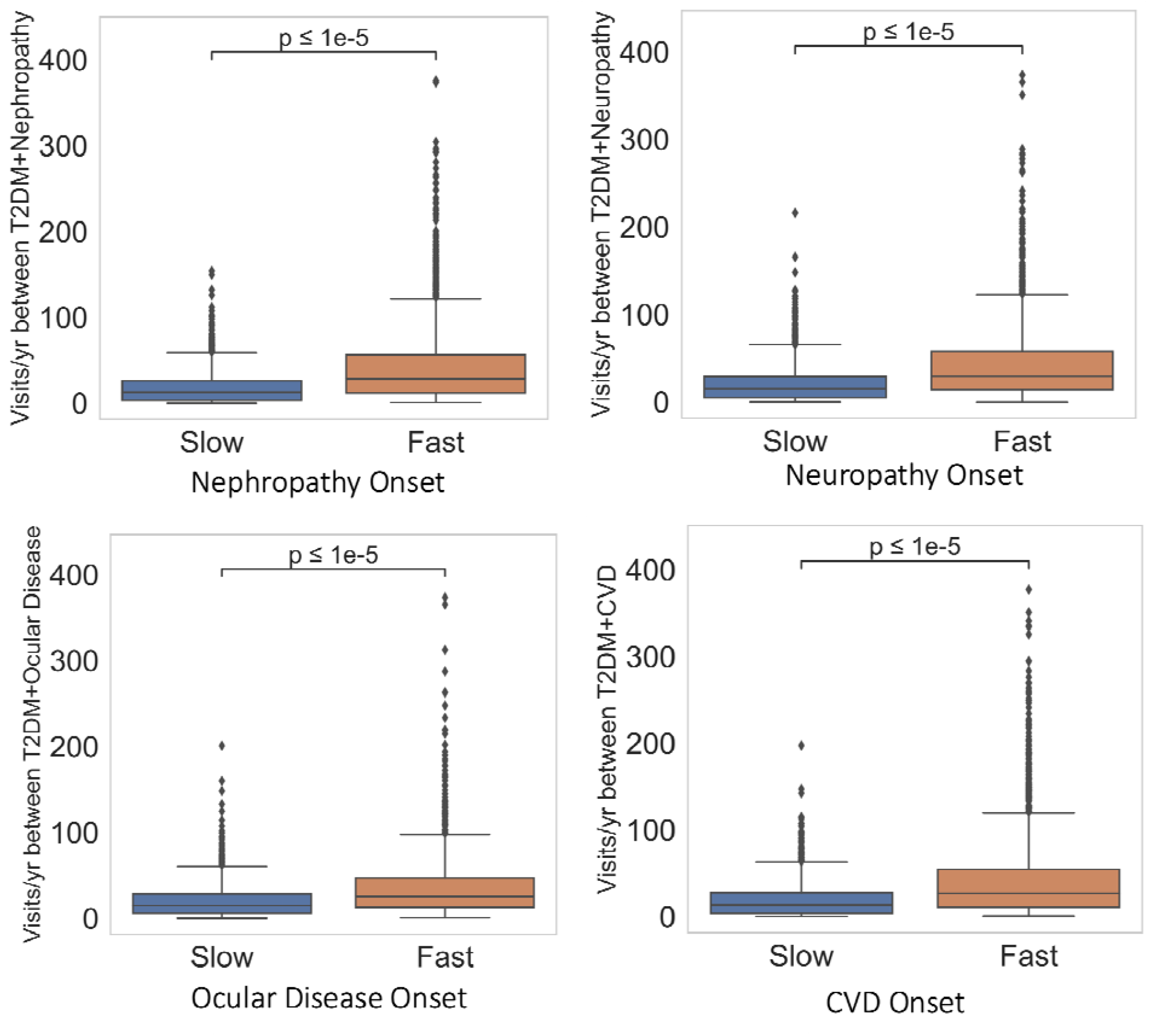
Boxplots comparing number of patient medical visits per year between T2DM and complication diagnoses between fast and slow complication onset group. Horizontal line within each box represents median and the box spans the interquartile range(IQR), extending from the 1^st^ (Q1) to the 3^rd^ (Q3) quartile for each group’s distribution. Box whiskers denote maximum (Q3+1.5*IQR) and minimum(Q1-1.5*IQR); dots outside of whiskers are outliers. Horizontal bar denotes p-value using two-sided Mann-Whitney-U test.

**Figure 6.**
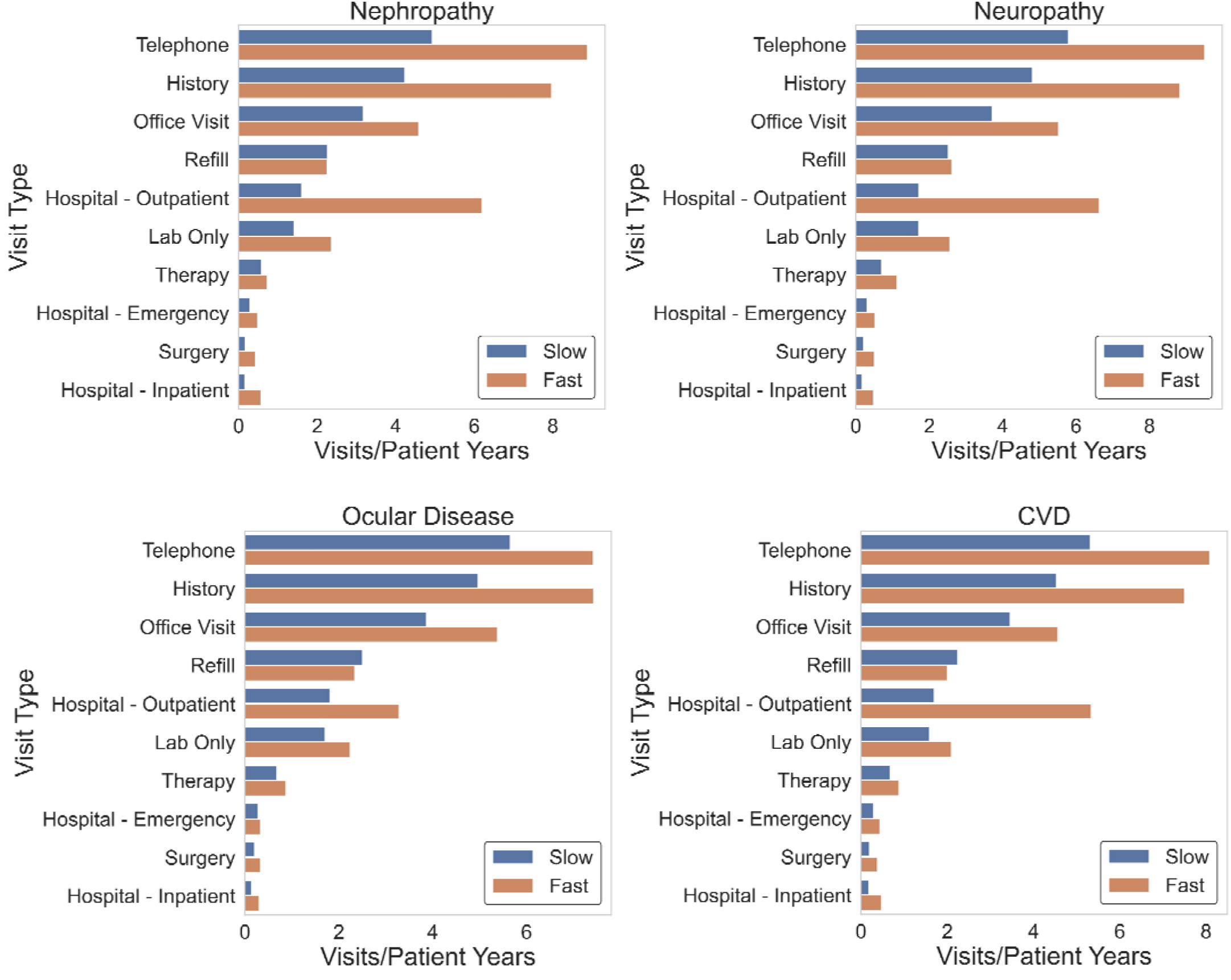
Barchart exhibiting types of medical visit obtained by fast and slow complication onset groups between T2DM and complication diagnoses. Number of different medical visits per patient year was visualized to assess differences in level of care obtained between groups. Patient year was defined as the sum of the individual times to complication within each group.

## Discussion

Our study developed well-calibrated models that can predict the development of a progressive diabetic complication (neuropathy, nephropathy, CVD, or ocular disease) before or after the median time to onset for each complication. The models performed best in distinguishing between fast and slow onset of nephropathy (AUROC 0.75) and worst in distinguishing fast and slow onset of CVD (0.70). One strength of the study was the ability of our model to perform with acceptable predictive performance using a smaller cohort relative to similar studies and traditional ML algorithms, which may be more easily implemented in clinical practice requiring less data than deep learning methods.

Evaluation of each data sources’ predictive performance allowed for understanding the utility of each data source in predicting onset of different diabetic complications. The combination of all five data sources (vitals, demographics, phenotypes, laboratory, and social history) was best. Use of social history or laboratory values alone as inputs led to the highest model performance. Laboratory values were most useful in predicting onset of ocular disease and CVD, social history was most useful for predicting onset of neuropathy, and laboratory and social history contributed equally to prediction of onset of nephropathy.

Unfortunately, there is lack of objective definitions of what makes a ML model ‘interpretable’ in clinical practice, and few works evaluate model usability for clinicians [30][31]. Well-established associations identified were: 1) lower eGFR (or reduced kidney function) was linked to faster onset of nephropathy, 2) higher anion gap (or increase in ketoacids in uncontrolled diabetes[32]) was linked to faster onset of all four complications, and 3) hyperlipidemia (an established risk factor for diabetic retinopathy[16]) was linked to faster onset of ocular disease.

Other less well-established findings from the SHAP analysis prompted further investigation. We were able to verify several predictors of diabetic complications identified by SHAP with the existing literature, although studies were often retrospective and included small patient cohorts.

Smokeless tobacco use status, a social history variable, was unexpectedly the most important feature across complications. In our data, smokeless tobacco refers to chew and snuff. Individuals with unknown smokeless tobacco use had a slower onset of any diabetic complication, and individuals who reported never using smokeless tobacco had faster onset of any complication. Across complications, the majority of patients in both groups were marked as having unknown smokeless tobacco use; when tobacco use was recorded, the percentages of cigarette and current smokeless tobacco users were higher among the fast onset complication group **(Tables 1-4)**. It is widely accepted that cigarette smoking accelerates vascular damage and increases the risk of cardiovascular morbidity/mortality in patients with T2DM, however the link to microvascular complications is not as clearly defined in diabetics[33]. As the progression of complications varies widely between smokers with T2DM, phenotypic predictors of susceptibility in diabetics to the negative effects of smoking warrants further investigation[33]. Prevalence rates of smoking in diabetics are similar to those of the general population, indicating diabetics continue to smoke despite the well-known health risks[33]. Alternative strategies that lead to risk reduction include use of smokeless tobacco and electronic cigarettes, which have been shown to be less harmful than combusted cigarettes and more effective options in helping smokers quit compared to nicotine-replacement products, although their long-term effects are unknown[34][35]. Thus, our findings that “never users” of smokeless tobacco had faster onset of complications may indicate a potential link between use of alternative smoking cessation strategies and slowing of diabetic complication progression.

Several laboratory values identified through SHAP had high importance in complication onset prediction but are non-traditional risk factors for diabetic complications. These may serve as new simple, cost-effective biomarkers for monitoring and prevention of diabetic complications, and further research is needed on reversibility of the complication with correction of the laboratory value.

First, higher MPV was associated with faster onset of nephropathy, neuropathy, and CVD in our study. A higher MPV is indicative of larger, younger, and more aggregable platelets that produce more pro-coagulants, such as thromboxane A2[36]. This platelet activation contributes to thrombogenesis, atherosclerosis, and production of oxidative substances like platelet-derived growth factor (PDGF) and vascular endothelial growth factor (VEGF) that cause local vascular lesions[36][37]. Small retrospective studies have shown that MPV and percentage of those developing diabetic complications were higher in patients with uncontrolled T2DM (HbA1c >7%) compared to those with controlled T2DM (HbA1c</=7%)[38,39]. Furthermore, improved glycemic control led to recovery in platelet activity, indicating the possibility of prevention of damaging platelet effects[40]. Overall, high MPV is associated with vascular damage in diabetics, and we may be able to prevent this damage through optimizing blood glucose control.

Second, individuals with lower bilirubin, lower MCHC and higher serum calcium had faster onset of ocular disease in our study. High levels of bilirubin, a breakdown product of hemoglobin, may indicate liver damage[41]. However, bilirubin may also have the potential to suppress oxidation of lipids and lipoproteins, a protective property against development of diabetic complications[42]. Several studies, including a meta-analysis of 27 studies, have shown low levels of bilirubin were inversely related to the development of diabetic complications, including retinopathy[42][43][44][45]. Next, several observational studies have shown that low hemoglobin levels may accelerate microvascular damage in diabetes. Low hemoglobin concentrations are more common in diabetic patients than non-diabetics and hyperglycemia has been shown to decrease red cell survival by 13%[46]. Studies have found an increased risk of severe diabetic retinopathy in individuals with hemoglobin levels below 12 g/dL[47][48], although this association diminished after adjusting for diabetes duration in another study[46]. Lastly, a cross-sectional study of over 3,000 patients found elevated serum calcium to be a risk factor for vision-threatening diabetic retinopathy[49], and *in vivo* histology of the retina revealed elevated serum calcium was associated with retinal photoreceptor apoptosis in diabetic retinopathy[50]. Low bilirubin and MCHC and high serum calcium in T2DM may be indicators of accelerated retinal damage in diabetics, providing clinicians with more personalized information for monitoring and modulating diabetes complication progression.

Third, our findings that higher RDW and lower MCHC are associated with faster onset of nephropathy are also supported by existing studies. RDW, which measures the volume and size of red blood cells, is commonly used to help diagnose different types of anemia[51]. A retrospective study of individuals with biopsy-proven diabetic nephropathy showed that individuals with higher RDW had an increased risk of progression to end-stage renal disease[51]. Diabetic patients with low hemoglobin concentration had more rapid decline in glomerular filtration[52], and anemia was a risk factor for progression to end stage renal failure[53]. High RDW and low MCHC may be important markers for progression of kidney injury in diabetics.

Of the demographic variables, only one, patient age at the time of diabetes diagnosis, was a top 10 predictor. Older individuals had faster onset of diabetic complications, which may be explained by reduced end organ reserve due to aging and comorbidities leading to faster organ damage in the elderly[16].

Other features we identified through SHAP require further investigation in defining their relationship to diabetic complication progression. For example, our study showed a higher monocyte percentage was associated with faster onset of ocular disease. Limited and contradictory evidence exists regarding the role of elevated monocyte counts and their effects in retinal cells of diabetics[54][55]. We also found lower BMI and pulse were associated with faster onset of CVD and answering “never” or “no” to illicit drug use was associated with faster onset of neuropathy or CVD, respectively. Lastly, body temperature was positively associated with faster onset neuropathy or ocular disease in our study. Diabetes is associated with reduced ability to dissipate heat during thermal stress, however minimal research exists that evaluates the effect of thermoregulatory control and long-term consequences in diabetics[56]. These are unexpected but potentially impactful findings that require further research.

Lastly, we explored follow-up care sought between the time of T2DM and complication diagnosis as a potential opportunity for improved diabetes care in the fast-onset group. However, we found that the fast-onset group had more medical visits (approximately biweekly compared to monthly in the slow-onset group) and the majority of visits were within the ambulatory setting. The Centers for Disease Control and Prevention (CDC) guidelines recommend a doctor visit and HgbA1c every 3 months if diabetes treatment goals are not being met, and every 6 months if goals are met[57]. As outpatient diabetes care may already be maximized, the focus may need to be switched to prevention of complication onset and the need for non-traditional, more personalized strategies.

This study has important limitations. First, data was collected from facilities across a single health network, so it is possible the models focused on features that are not as common or important in other institutions. The models should be implemented on a larger scale across different institutions to verify reproducibility. Second, data may not be reliably recorded in patient electronic records. For example, across complications, the majority of patients had ‘unknown’ smokeless tobacco use. Asking about smokeless tobacco use may not be standard history-taking practice, and practice may vary across different facilities across the health network. Third, certain clinically relevant variables were excluded due to high missingness in the dataset (>50%), such as HgbA1c. Lastly, further research is warranted to integrate other clinical risk indicators, such as medications, imaging and patient-specific proteomics data to create a more complete prediction model.

In this study, ML models are able to accurately predict the onset of one of diabetic complications: neuropathy, nephropathy, ocular disease and CVD. SHAP provides an interpretation of key features’ contribution to each model, allowing clinicians to understand which patient markers place individuals at high risk of fast progression to a complication at the time of their T2DM diagnosis. Our study is unique in the realm of ML studies as it explores the relationship between patient biomarkers not routinely used in diabetes monitoring, such as bilirubin, calcium, and MPV, and onset to diabetic complications. These markers may serve as economical tests for more tailored monitoring and prevention of progression to a diabetic complication, and larger, more robust studies are needed to investigate these associations. In conclusion, a combination of ML and SHAP can serve as a starting point for better prediction and understanding of disease risk.

## Methods

### Study population

This was a retrospective study across an academic hospital network to predict rapid versus delayed onset of diabetic complications in individuals with T2DM. Retrospective, de-identified patient data was queried using the Medical College of Wisconsin (MCW) Clinical Research Data Warehouse using the Froedtert Health System’s Informatics for Integrating Biology and the Bedside (i2b2) tool and extracted using the Froedtert Health System Honest Broker. 30,854 patients were identified who had a diagnosis code for T2DM followed by at least one of our complication codes. Extracted data spanned over 24 years from May 1997 to August 2021.

### Data collection

T2DM diagnosis was defined as the date of the first ICD-9 code of 250.00 (T2DM without complications) or ICD-10 code of E11.9 (T2DM without complications). The US Department of Health and Human Services required the US transition to the use of only ICD-10 codes in October of 2015[58], thus an individual diagnosed prior to 2015 would be coded with 250.00; if the diagnosis occurred after 2015, an E11.9 would have been coded. In order to exclude individuals who had an occurrence of a diabetic complication prior to their first T2DM diagnosis, a temporal query was used to extract individuals who had a diagnosis of T2DM without complications (250.00 or E11.9) that occurred prior to a diagnosis of one of four complications[25][59]:

⍰ Nephropathy: 250.40, 403, 404, 581, 583, 584, 585, 586, 588, 593, E11.2, I12, I13, N04, N05, N08, N17, N18, N19, N25, N29
⍰ CVD: 250.70, 410, 412, 413, 414, 428, E11.5, I20, I21, I25, I50
⍰ Ocular Diseases: 250.50, 362, 365, 366.41, E11.3, H35
⍰ Neuropathy: 250.60, 337.1, 353.5, 354.8, 354.9, 355.7, 355.8, 357.2, E11.4, G62.9, G63.

Cerebrovascular disease was not included as a complication due to limited data (750 instances). Patients who had a time-to-complication less than one month were excluded from the study to avoid inclusion of patients who had diabetic complications diagnosed at the same time as their T2DM diagnosis. Per the American Diabetes Association (ADA) guidelines, a one month follow-up visit is advised for diabetes care for all patients with hyperglycemia in the inpatient setting; thus, if a patient was diagnosed with T2DM for the first time in the hospital, a follow-up and assessment of whether complications were present within one month is considered standard of practice[60]. After excluding these individuals, the number of individuals who had a diagnosis code for diabetes followed by at least one complication code decreased to 21,850 patients (**Figure 1A**).

The study included data from multiple sources, including demographic information, laboratory results, social-lifestyle history, vital signs, and ICD-9/10 diagnosis codes. The data were linked using de-identified unique encoded patient numbers. Only ICD codes before or on the date of T2DM diagnosis were used as model inputs. Diagnosis codes included both ICD-9 and ICD-10 versions. To unify across all patients, codes were replaced with the corresponding phenotype within the phecode system[61][62]. This also helped to prevent the model learning unintended associations linked to the longer existence of an ICD-9 versus an ICD-10 code rather than the code itself. Phecodes are distinct diseases or traits that map to ICD-9 or ICD-10 codes as a means to provide consistency across these codes over time as well as overlapping disease states [63]. For example, 401.1 (ICD-9) and I10 (ICD-10) would both map to the phenotype ‘Essential hypertension’[26, 27]. 30,854 unique diagnosis codes were converted to 1,721 unique phenotypes.

Demographics information comprised of seven input features: sex, age, ethnicity, race, employment status, marital status, and language. Demographics information did not change over time.

Because patients had many entries for vitals, social-lifestyle history, and laboratory values, and to simulate how our models might be used in the real world, we used the last collected data before or on the day of the date of T2DM diagnosis. Vital signs included body mass index, diastolic blood pressure, systolic blood pressure, pulse, temperature and respiration rate. Social-lifestyle history consisted of alcohol use, illicit drug use, tobacco use (cigarettes, pipes, and cigars) and smokeless tobacco use (snuff and chew). Laboratory values consisted of aspartate aminotransferase (AST), alanine transaminase (ALT), bilirubin, alkaline phosphatase, calcium, glucose, bicarbonate, chloride, sodium, potassium, creatinine, eGFR, eGFR for African Americans, blood urea nitrogen, anion gap, platelet count, hematocrit, hemoglobin, red blood cell count, white blood cell count, MCHC, mean corpuscular volume, MPV, RDW, monocyte percentage, neutrophil percentage, eosinophil percentage, lymphocyte percentage, absolute neutrophil count, absolute lymphocyte count, absolute monocyte count, absolute eosinophil count, total protein, and albumin. HgbA1c was not included as a laboratory parameter since there was >50% missingness for this value in the dataset.

### Data preprocessing

Features with more than 50% of values missing were excluded. MissForest imputation was used to impute missing values[64]. Input data was filtered to only include values collected on a visit occurring the day of or prior to the initial T2DM diagnosis in order to mirror a clinical scenario where a clinician only has access to the patient’s baseline health records at the time of T2DM diagnosis. Categorical variables were one-hot encoded[65], continuous variables were normalized using Min-Max normalization, and counts of phenotypes for each column were binarized (**Supplementary Figure 6**). Any values +/-3 standard deviations from the mean for a particular feature were set to N/A and then imputed because these values are likely reporting errors.

Continuous patient baseline variables were reported as the median (inter-quartile range) and cohort differences were tested using a two-sided Mann-Whitney-U test. Categorical variables were reported as counts (percentages) and compared using chi-square test. Statistical significance was based on a two-tailed p-value of ≤ 0.05.

### Study outcomes

The primary endpoint of the study was classification of a diabetic complication (neuropathy, nephropathy, CVD, or ocular disease) prior to or after the median time to complication (years). Individuals who developed a complication prior to the median time were classified as having fast onset of a complication, those with a time to complication longer than the median were classified as having slow onset. Using the median time as the cut-off between the two groups allowed for balanced classification.

### Machine Learning

Six supervised machine learning methods were trained to generate a prediction model for onset of diabetic complications, including GB, SVC, RF, ET, LR, and AdaBoost. Each model was optimized separately to predict the four complications with one of six input datasets: phenotypes, demographics, vital signs, social-lifestyle history, laboratory data, and all inputs combined. A total of six models were optimized with the six potential inputs for each of the four complications, resulting in a total of 144 model/input/output combinations that were optimized.

Data was split into 20% final test and 80% training data. The 80% training split was used to tune hyperparameters; each model was evaluated using a randomized search (RandomizedSearchCV) with 5-fold cross validation, up to 1,000 iterations, and AUROC metric used for scoring. The hyperparameters that maximized the average AUROC values obtained from the randomized search were used to refit a model on the 80% training dataset, and the test set was used to evaluate generalization performance of the best model. Best hyperparameters corresponding to the best model for each input are shown in **Supplementary Table 1**. AUROC scores reported in this study represent performance of the test set. The input and corresponding model with the best performance for each complication were calibrated via parametric ‘sigmoid’ method and 5-fold cross validation of the CalibratedClassifierCV class. Model calibration was assessed by plotting calibration curves of the observed versus predicted probabilities for the positive class across 10 evenly partitioned bins. The brier scores for each calibration plot was calculated using the true class values and the predicted probabilities of the test set.

### Model interpretation

SHAP[21] [66] values were used identify features that contribute most to model prediction. For consistency, the random forest classifier models with all data as input were used for SHAP analysis of each complication.

To better understand the relationship between number of medical visits and time of complication diagnosis, we used the patient encounters database to derive individuals’ inpatient and outpatient visits between their T2DM and complication diagnoses. Number of visits per year between slow and fast diagnosis groups were compared using a two-sided Mann-Whitney-U test. Statistical significance was based on a two-tailed p-value of ≤ 0.05. The number of each type of medical visit between the T2DM and complication diagnoses divided by patient years was further visualized to assess level of care obtained. Patient years, defined as the sum of the individual patient complication times in each group, was used to account for differences in the total years of follow-up between fast and slow complication onset groups.

### Statistical analysis

All data cleaning, analysis, and model training were performed in Python version 3.7.11 (Scikit-Learn[67], SciPy[68], SHAP[21]) and R (MissForest[64]).

## Data Availability

All data produced are available online at https://github.com/amandamomenzadeh/ML-Predict-T2DM-Comp-Onset

## Code availability

All code is available from github https://github.com/amandamomenzadeh/ML-Predict-T2DM-Comp-Onset

## Acknowledgements

This work was supported by startup funds from the Medical College of Wisconsin, the National Center for Research Resources, and the National Center for Advancing Translational Sciences, National Institutes of Health, through Grant Number UL1TR001436. Its contents are solely the responsibility of the authors and do not necessarily represent the official views of the NIH. This research was completed in part with computational resources and technical support provided by the Research Computing Center at the Medical College of Wisconsin. We thank Eduard Puig for graphic design help.

## Author contributions

Conceptualization, A.M., A.S., J.G.M; Methodology, A.M., A.S., J.G.M.; Software, A.M., A.S., J.G.M.; Formal Analysis, A.M., A.S., J.G.M.; Data Curation, A.M., A.S., J.G.M.; Writing – Original Draft, A.M., A.S., J.G.M.; Writing – Review & Editing, A.M., A.S., J.G.M.; Visualization, A.M., A.S., J.G.M.; Supervision, J.G.M.; Project Administration, J.G.M.; Funding Acquisition, J.G.M.

## Materials & Correspondence

Amanda Momenzadeh

