## Supplemental Figures for "Clinical interpretation of machine learning models for prediction of diabetic complications using electronic health records"

**Supplementary information**

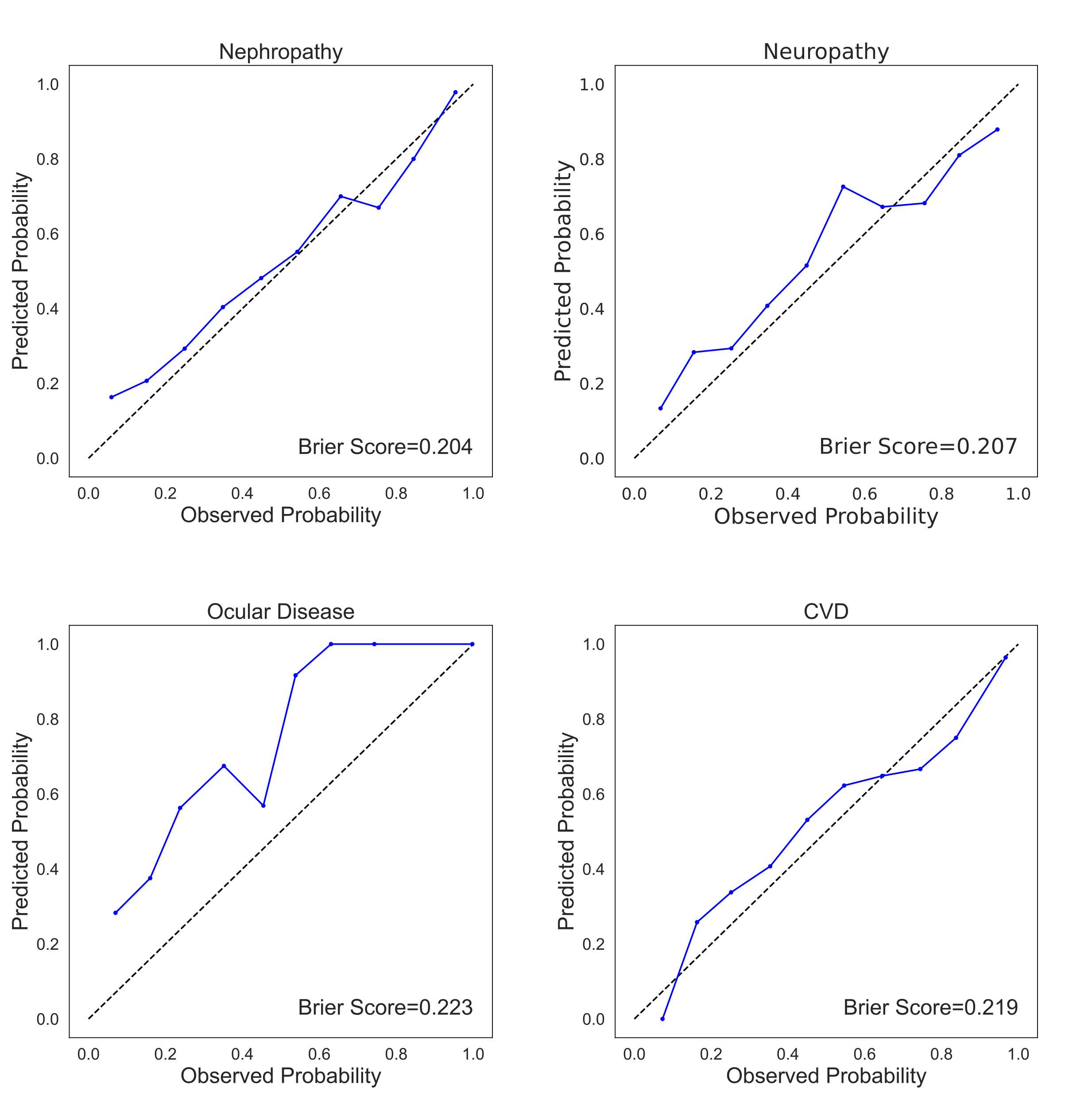

**Supplementary Figure 1 Calibration Plots.** For each complication, the model with the best performance using all inputs combined sas calibrated via the parametric ‘sigmoid’ method and 5-fold cross validation of the CalibratedClassifierCV class. Each calibration curve plots the observed versus predicted probabilities for the positive class across 10 evenly partitioned bins. The brier scores for each plot was calculated using the true class values and the predicted probabilities of the test set.

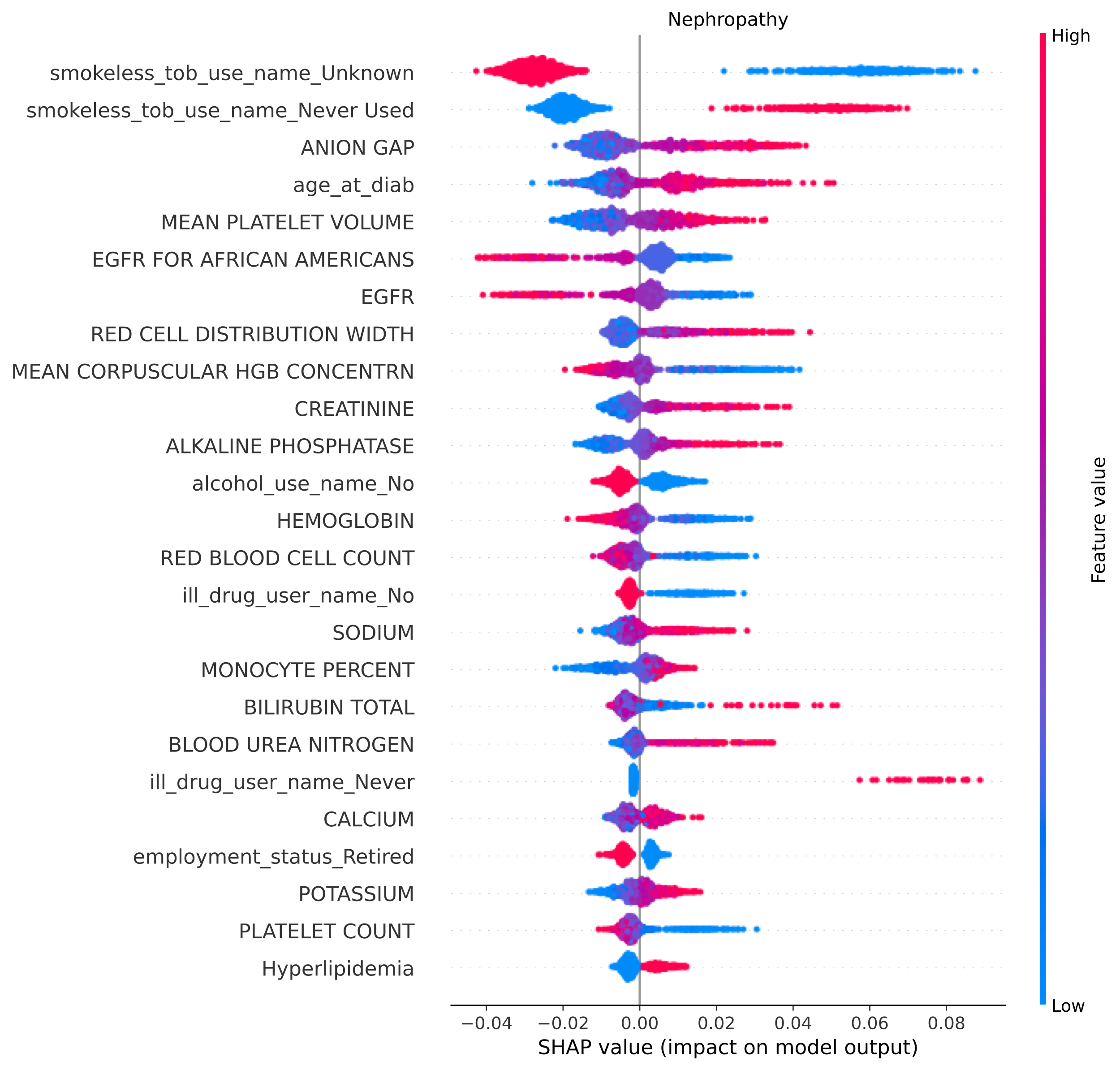
**Supplementary Figure 2 Top 25 features for Nephropathy visualized using SHAP.** Individual patient contributions to the outcome are signified with red dots (high feature values), purple (intermediate), and blue (low). Y-axis represents importance of each feature. Dots with x values greater than and less than zero represent patients with a fast and slow complication onsets, respectively.

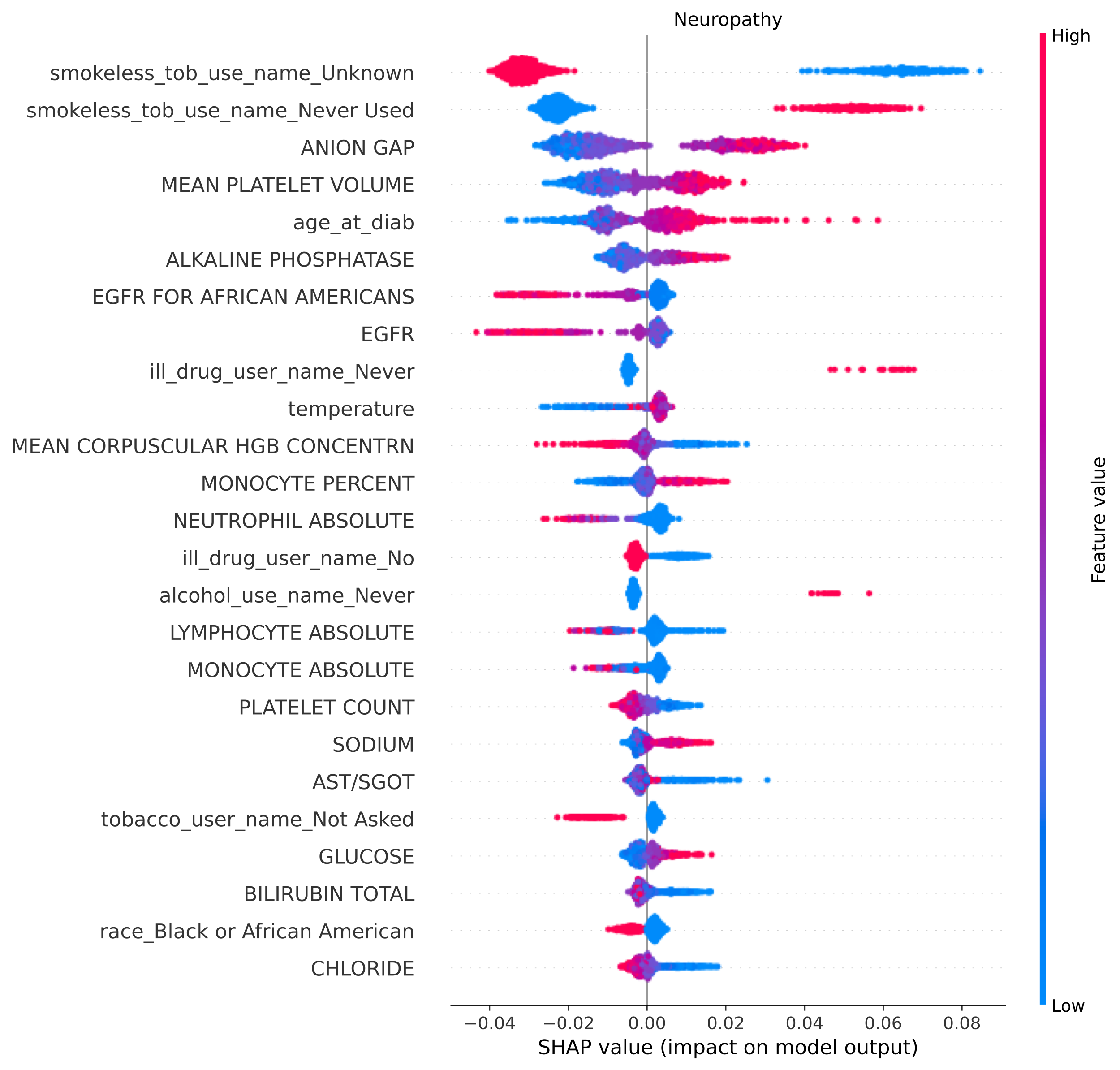

**Supplementary Figure 3 Top 25 features for Neuropathy visualized using SHAP.** Individual patient contributions to the outcome are signified with red dots (high feature values), purple (intermediate), and blue (low). Y-axis represents importance of each feature. Dots with x values greater than and less than zero represent patients with a fast and slow complication onsets, respectively.

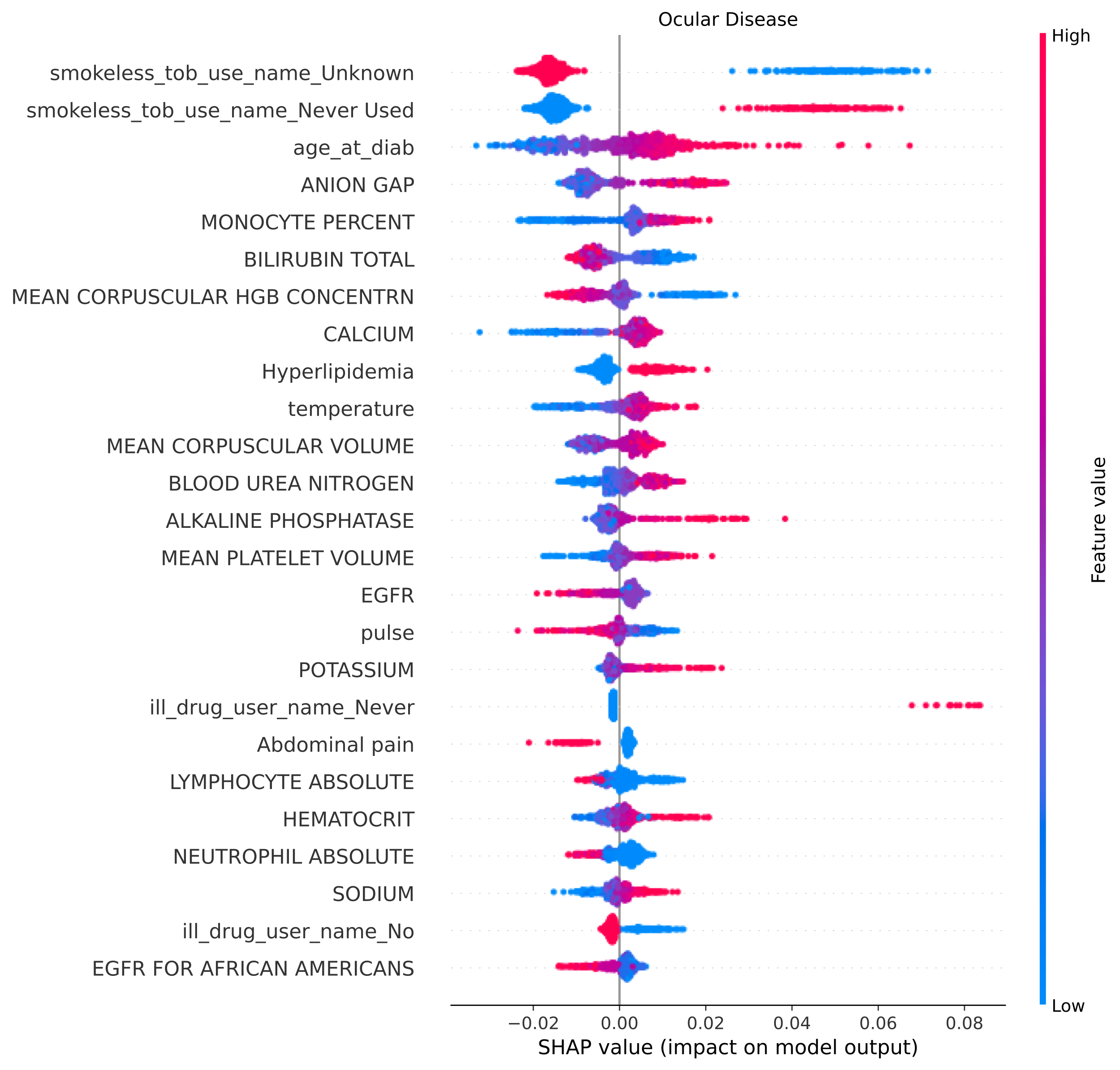
**Supplementary Figure 4 Top 25 features for Ocular Disease visualized using SHAP.** Individual patient contributions to the outcome are signified with red dots (high feature values), purple (intermediate), and blue (low). Y-axis represents importance of each feature. Dots with x values greater than and less than zero represent patients with a fast and slow complication onsets, respectively.

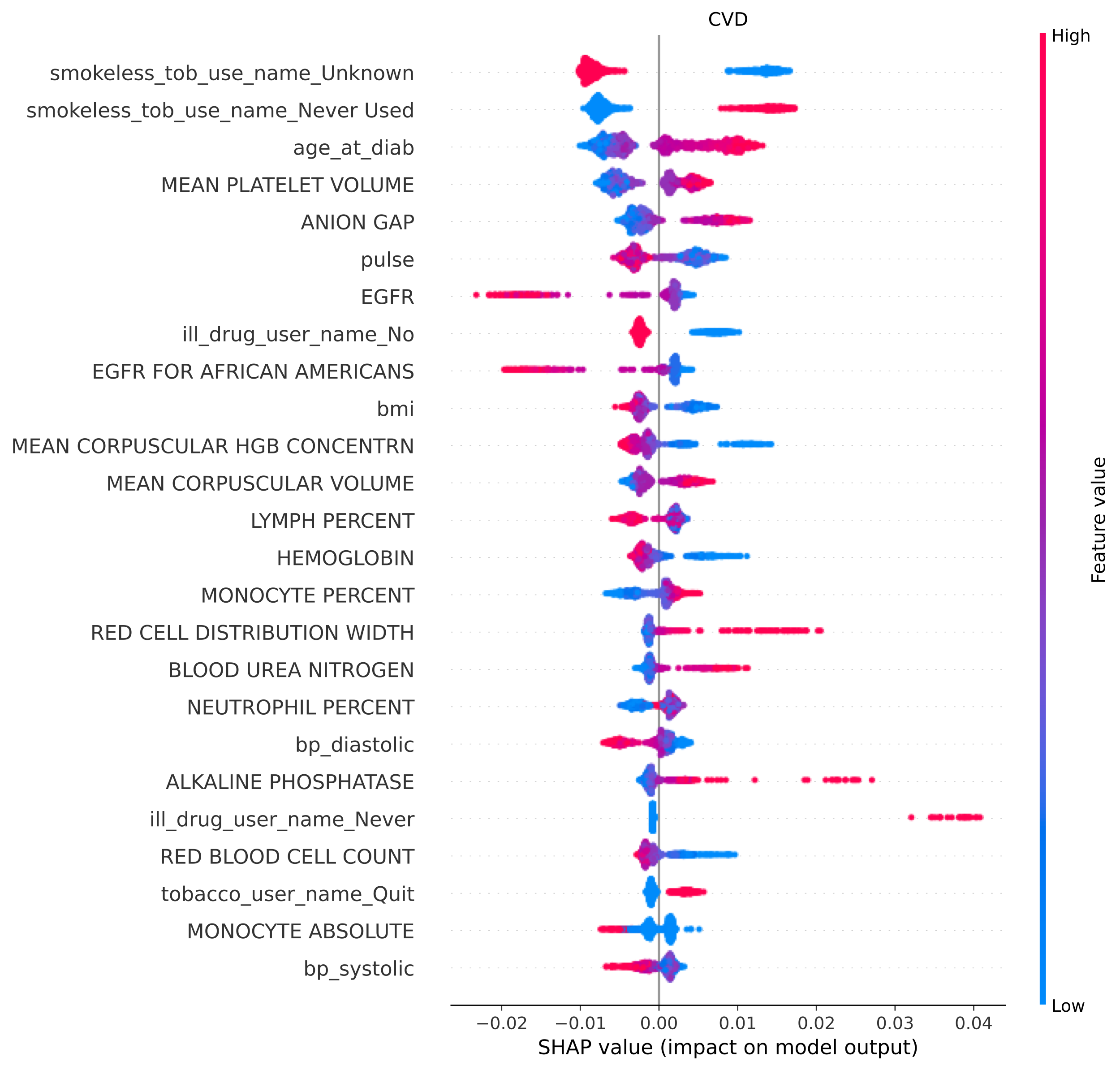
**Supplementary Figure 5 Top 25 features for CVD visualized using SHAP.** Individual patient contributions to the outcome are signified with red dots (high feature values), purple (intermediate), and blue (low). Y-axis represents importance of each feature. Dots with x values greater than and less than zero represent patients with a fast and slow complication onsets, respectively.

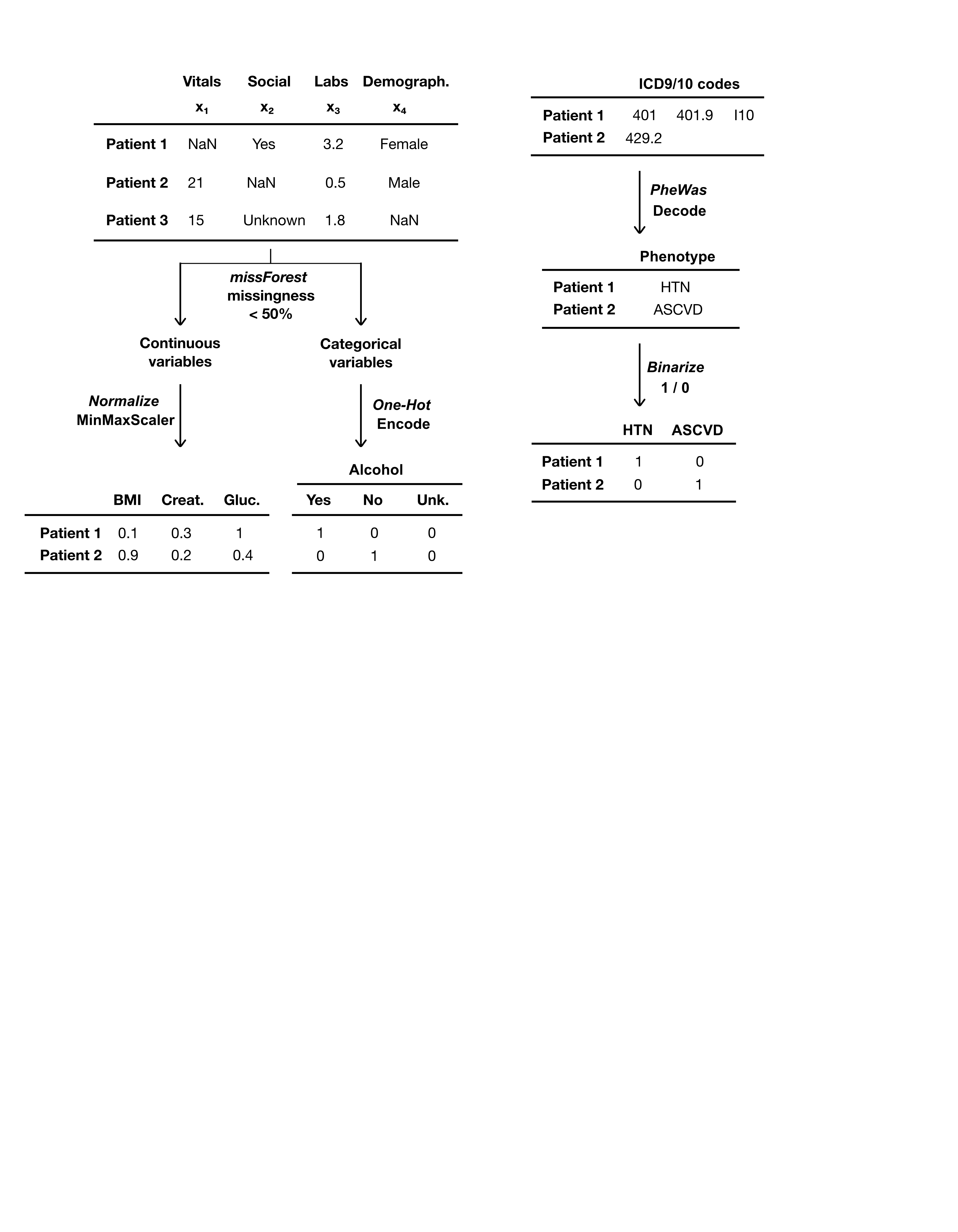

**Supplementary Figure 6 Data Pre-processing.** For vitals, social-lifestyle history, laboratory, and demographics inputs, MissForest was used to impute missing values for features with less than 50% missingness. Continuous variables were normalized using Min-Max normalization and categorical variables were one-hot encoded. ICD9/10 codes were converted to phenotypes using the PheWas system and counts of phenotypes for each column were binarized.
