## Supplemental Table for "Clinical interpretation of machine learning models for prediction of diabetic complications using electronic health records"

| **Complication** | **Input** | **Best Model** | **Best Hyperparameters** |
| --- | --- | --- | --- |
| **Nephropathy** | Phenotypes  Demographics  Vitals  Social  Labs  All | ET  AdaBoost  RF  ET  RF  RF | bootstrap=True, max_depth=100.0, min_samples_leaf=2, min_samples_split=10,  n_estimators=1000  learning_rate=0.45  max_depth=45.0, min_samples_leaf=150, min_samples_split=80, n_estimators=50  bootstrap=True, max_depth=70.0, min_samples_leaf=10, min_samples_split=450,  n_estimators=10  max_depth=40.0, max_features='log2', min_samp;es_leaf=10, n_estimators=500  bootstrap=False, max_depth=95.0, max_features ='sqrt', min_samples_leaf=2,  n_estimators=2000 |
| **Neuropathy** | Phenotypes  Demographics  Vitals  Social  Labs  All | RF  AdaBoost  AdaBoost  ET  RF  RF | max_depth=90.0, max_features='sqrt', min_samples_leaf=2, min_samples_split=100,  n_estimators=250  learning_rate=0.6  learning_rate=0.55, n_estimators=10  max_depth=5.0, max_features='log2', min_samples_leaf=2, min_samples_split=500,  n_estimators=500  max_depth=20.0, max_features='log2', min_samples_leaf=20, min_samples_split=100  bootstrap=False, max_depth=70.0, min_samples_leaf=2, min_samples_split=150,  n_estimators=250 |
| **Ocular Disease** | Phenotypes  Demographics  Vitals  Social  Labs  All | SVC  LR  RF  SVC  LR  AdaBoost | C=10, gamma=0.001, probability=True  C=0.01, penalty='none', solver='sag'  bootstrap=False, max_depth=95.0, max_features='log2', min_samples_leaf=20,  min_samples_split=400, n_estimators=10  C=1, gamma=0.1, probability=True  C=0.01, penalty='none', solver='sag'  learning_rate=0.15 |
| **CVD** | Phenotypes  Demographics  Vitals  Social  Labs  All | RF  GB  ET  ET  SVC  ET | max_depth=85.0, max_features='sqrt', min_samples_leaf=2, min_samples_split=200,  n_estimators=500  learning_rate=0.2, max_depth=70.0, max_features='auto', min_samples_leaf=250,  min_samples_split=350, n_estimators=10, subsample=1  max_depth=95.0, max_features='sqrt', min_samples_leaf=10, min_samples_split=10,  n_estimators=250  bootstrap=True, max_depth=60.0, max_features='sqrt', min_samples_leaf=10,  min_samples_split=500, n_estimators=50  C=1, gamma=0.1, kernel='poly', probability=True  bootstrap=True, max_depth=95.0, min_samples_leaf=2, min_samples_split=10,  n_estimators=500 |

**Supplementary Table 1 Best Hyperparameters.** Hyperparameters optimizing test dataset AUROC corresponding to the best model for each input.
